# Temporal trends and clinical characteristics of acute hepatitis with unknown aetiology and human adenovirus infections in Oxfordshire from 2016 to 2022

**DOI:** 10.1101/2023.06.19.23291626

**Authors:** Cedric CS Tan, Gavin Kelly, Jack Cregan, Joseph D Wilson, Tim James, Meera Chand, Susan Hopkins, Maaike Swets, J Kenneth Baillie, Katie Jeffery, Sarah Walker, David W Eyre, Nicole Stoesser, Philippa C Matthews

**Affiliations:** The Francis Crick Institute, London, UK; UCL Genetics Institute, University College London, London, UK; Bioinformatics and Biostatistics, The Francis Crick Institute, London, UK; Nuffield Department of Medicine, University of Oxford, Oxford, UK; King’s College Hospital NHS Foundation Trust, London, UK; Department of Biochemistry, Oxford University Hospitals NHS Foundation Trust, Oxford, UK; United Kingdom Health Security Agency, Colindale, UK; NIHR Health Protection Research Unit, Imperial College London, London, UK; Leiden University Medical Center, Leiden, Netherlands; Roslin Institute, University of Edinburgh, Edinburgh, UK; Department of Infectious Diseases and Microbiology, Oxford University Hospitals, Oxford, UK; Radcliffe Department of Medicine, University of Oxford, Oxford, UK; Oxford University Hospitals NHS Foundation Trust, Oxford, UK; Big Data Institute, Nuffield Department of Population Health, University of Oxford, Oxford, UK; University College London Hospitals NHS Foundation Trust, London, UK; University College London, London, UK

**Keywords:** hepatitis, acute, adenovirus, outbreak, AAV, epidemiology, electronic health records, surveillance

## Abstract

**Background:** An outbreak of severe acute hepatitis of unknown aetiology (AS-Hep-UA) in children during 2022 has subsequently been linked to infections by adenovirus-associated virus 2 (AAV2) and other ‘helper viruses’, including human adenovirus (HAdV).

**Aim:** We investigated clinical characteristics and temporal distribution of acute hepatitis with unknown aetiology (AHUA) and of HAdV infections in Oxfordshire, UK population between 2016-2022.

**Methods:** We used anonymised electronic health records (EHR) to collate retrospective data for presentations of AHUA and/or HAdV infection between 2016-2022. We reviewed records of >900,000 acute presentations to emergency care at Oxford University Hospitals NHS Foundation Trust (OUH; UK) and performed a descriptive analysis of case numbers and clinical characteristics.

**Results:** Over the full study period, patients coded as AHUA had significantly higher critical care admission rates (p<0.0001, OR=41.7, 95% CI:26.3-65.0) and longer inpatient admissions (p<0.0001) compared with the rest of the patient population. Comparing events within the outbreak period (1st Oct 2021 - 31 Aug 2022), to those occurring outside this period, significantly more adults were diagnosed with AHUA (p<0.0001, OR=3.01, 95% CI: 2.20-4.12), and there were significantly more HAdV infections in children (p<0.001, OR=1.78, 95% CI:1.27-2.47). There were also more HAdV tests administered during the outbreak (p<0.0001, OR=1.27, 95% CI:1.17-1.37). There was no evidence of more acute hepatitis or increased severity of illness among patients who tested HAdV-positive compared to those testing HAdV-negative.

**Conclusion:** Our results highlight an increase in the number of AHUA in adults coinciding with the reported AS-Hep-UA outbreak in children, but not linked to documented HAdV infection.

## INTRODUCTION

In April 2022, the United Kingdom Health Security Agency (UKHSA) alerted the World Health Organization (WHO) to a significant increase in acute severe hepatitis in children aged less than 10 years, who were otherwise clinically fit and well, dating back to January 1st 2022 (1). Concerningly, a proportion of these children had sufficiently severe disease to warrant liver transplantation (2). Initial investigations and evaluation demonstrated no link to Hepatitis viruses A-E, other known causes of acute hepatitis, toxins, common exposures, or foreign travel; these cases were therefore designated ‘acute severe hepatitis of unknown aetiology’ (AS-Hep-UA).

Subsequent detailed investigation of samples from affected children suggested a likely infectious aetiology, with metagenomic sequencing identifying adeno-associated virus 2 (AAV2) in 81-96% AS-Hep-UA patients (versus 4-7% in controls), alongside a higher than expected prevalence of human adenovirus (HAdV) (3–5). In addition to HAdV, a likely contribution was made by AAV coinfection with other ‘helper’ viruses including acute infections or reactivation of latent infections, particularly with Epstein-Barr Virus (EBV), human herpes-virus 6 (HHV6) and enteroviruses (3–5), and/or a contribution from superantigen-mediated immune activation (6). A significant enrichment of the Human Leucocyte Antigen (HLA) class II allele DRB1*04:01 has been identified among AS-Hep-UA cases compared to the background population, suggesting a specific immune susceptibility (3).

Following the initial reporting of AS-Hep-UA in Scotland, several cases were retrospectively identified in the United States dating back to October 2021 (6). By the start of July 2022, >1000 probable cases had been identified worldwide (7). The outbreak in Europe peaked between the end of March and early May of 2022 (week 12 to 18), and subsequently declined between May and August (6). The case definition of AS-Hep-UA was refined to include anyone under the age of 16 presenting no earlier than October 1st 2021 with an acute hepatitis and deranged serum liver enzymes (alanine transaminase (ALT) or aspartate transaminase (AST) >500 IU/L) which could not be accounted for by other causes (7).

Despite AS-Hep-UA being identified worldwide, geographical disparities in the incidence of cases and liver transplantation relative to baseline were evident; rates in the UK and across parts of Europe clearly exceeded expected averages, in contrast to no significant deviation from baseline across the United States, Brazil, India, and Japan (8,9). More than a quarter of global cases were identified in the UK, which has a 100-fold relative incidence rate compared to France, despite the countries being geographical neighbours of almost identical population sizes (10). However, the relative contribution of enhanced surveillance, population susceptibility, and circulation of any causative agent to these differing rates has remained unclear. Many patients diagnosed with acute hepatitis with unknown aetiology (AHUA), but not meeting the stringent case definitions (i.e. ALT/AST < 500IU/L; age >15 years), would not have been reported as AS-Hep-UA cases. Therefore, whether the AS-Hep-UA outbreak was the ‘tip of an iceberg’ of milder cases of AHUA in the population during that period is not known.

Routinely collected clinical data (e.g. patient diagnoses, liver enzyme and microbiology test results) in the form of electronic health records (EHRs) present an opportunity to investigate population epidemiology of AHUA. Furthermore, there is utility in considering whether these routine clinical laboratory parameters could be used as a surveillance tool at a population level, for example as a sentinel marker for circulation of an infectious trigger or for increased incidence of disease in the population that may be cause for concern.

In this study, we used hospital EHR data from Oxfordshire, UK, to explore trends in AHUA and HAdV infections before, during and after the period of the AS-Hep-UA outbreak. We addressed three specific aims: (i) to explore any changes in liver enzyme levels in adults and children presenting to hospital, (ii) to determine any changes in incidence of AHUA and HAdV infections in adults and children, and (iii) to identify any associations between AHUA and/or HAdV infection with patient outcomes.

## METHODS

### Data source

We analysed EHRs of children and adults presenting as an emergency to Oxford University Hospitals (OUH) NHS Foundation Trust, a large tertiary referral hospital in the South East of England, serving a population of ∼725,000. Data were accessed through an application to the Infections in Oxfordshire Research Database (IORD) (11), which has ethical approvals from the National Research Ethics Service South Central – Oxford C Research Ethics Committee (19/SC/0403), the Health Research Authority and the national Confidentiality Advisory Group (19/CAG/0144), including provision for use of pseudonymised routinely collected data without individual patient consent. Patients who choose to opt out of their data being used in research are not included in the study. The study was carried out in accordance with all relevant guidelines and regulations. The study sponsor was Oxford University Hospitals (OUH). All patients were assigned an anonymised ‘cluster ID’, with no identifying details handled by the research team. We recorded month/year of birth without the specific date of birth and avoided disaggregation into any category containing <5 individuals. Data were held within a password protected, encrypted database and accessed only by named investigators in accordance with NHS standards for data management and protection.

### Data collection and definitions

We reviewed data from 1^st^ March 2016 to 31^st^ December 2022 for all individuals aged 18 months and older presenting to the Emergency Department or acute medical/surgical assessment units at OUH. We recorded subsequent admission to hospital, admission to the Intensive Care Unit (ICU), duration of hospital admission, and mortality during the admission. Patients were stratified into three categories based on their ages upon presentation to OUH: younger children (<7 years), older children (7-15 years) and adults (≥16 years). Epochs were considered as pre-COVID-19 (1st March 2016 - 10 March 2020), COVID-19 pandemic period (11th March 2020 - 31st December 2022), and nested within the COVID-19 pandemic period, the AS-Hep-UA outbreak (1st Oct 2021 - 31 Aug 2022).

### Laboratory data

Laboratory data were generated by externally ISO accredited clinical biochemistry and microbiology laboratories in the OUH. Parameters collected are summarised in **Table 1**. Laboratory data were based on those routinely collected at OUH, where a request for ‘liver function tests’ (LFTs) prompts a clinical biochemistry profile of four tests, alanine transferase (ALT), alkaline phosphatase (ALP), bilirubin (BR) and albumin (Alb). Additional laboratory investigations were requested at the discretion of the clinical team, which include aspartate transaminase (AST), gamma glutamyl transferase (GGT), C-reactive protein (CRP) and full blood count (FBC). FBC analysis included a white blood cell count (WBC) and was undertaken using a Sysmex XN automated analyser (Sysmex UK Ltd., Milton Keynes, UK). Liver function and CRP tests were undertaken using standard methods on Abbott Architect c16000 analysers (Abbott Laboratories, Maidenhead, UK). Reference intervals for liver enzymes and inflammatory markers are provided in **Table 1**. Abnormalities in these biomarkers were classified based on the upper limit of normal (ULN) for all ages and both sexes – mild, moderate and severe derangement was defined as up to 2x, 2-5x and >5x ULN, respectively, with the exception of albumin, which was classified as deranged if levels were less than the lower limit of normal (LLN) of 32g/L.

**Table 1:** Characteristics of population of adults and children aged ≥18 months presenting as an emergency to Oxford University Hospitals NHS Foundation Trust (UK) between 2016 and 2022.

| Characteristic | | | Younger children<br>(18 months-6<br>years)<br>n=70,962 | Older children<br>(7-15 years)<br>n=86,217 | Adults<br>($>15$ years)<br>n=746,254 |
| --- | --- | --- | --- | --- | --- |
| Age in years at presentation<br>(median, IQR) |  |  | 3 (2-5) | 11 (9-13) | 53 (32-74) |
| Male sex (%) |  |  | 57.1 | 54.2 | 47.5 |
| Median index of multiple deprivation score* |  |  | 11.5 | 11.1 | 11.5 |
| Proportion admitted to hospital following<br>emergency presentation (%) |  |  | 17.0 | 13.2 | 34.3 |
| Proportion admitted to ICU (%) |  |  | 0.4 | 0.3 | 1.5 |
| Mortality during admission episode (%) |  |  | 0.01 | 0.01 | 0.41 |
| Admission duration in hours if admitted, median<br>(IQR) |  |  | 20 (12-42) | 24 (14-49) | 56 (22-160) |
|  | **Biomarker** | **Ref. Range** |  |  |  |
| **Liver**<br>**biomarkers**<br>**(median, IQR)** | ALT | 10-45 IU/L | 15 (12-21) | 14 (11-20) | 19 (13-30) |
|  | AST | 15-42 IU/L | 41 (31-144) | 33 (21-119) | 41 (22-110) |
| | Bilirubin | 0-21 μmol/L | 5 (3-7) | 7 (5-11) | 9 (6-14) |
|  | Albumin | 32-50 g/L | 38 (35-40) | 40 (38-43) | 37 (33-40) |
|  | ALP | 30-130 IU/L | 195 (160-237) | 181 (115-247) | 79 (64-101) |
|  | GGT | 15-40 IU/L | 17 (10-104) | 20 (13-62) | 85 (31-280) |
| **Infection**<br>**biomarkers**<br>**(median, IQR)** | CRP | 0-5 mg/L | 10 (1.3-42.3) | 2.5 (0.4-20.5) | 8.6 (2.1-43.4) |
| | WBC count | 4-11 $\times 10^9$/L | 10 (7.51-13.8) | 8.53 (6.58-11.5) | 8.87 (6.9-11.6) |

\* Measure of poverty based on multiple components such as employment, income, and disability.

HAdV testing was undertaken using a PCR-based multiplex test on respiratory samples (FilmArray Respiratory 2.1 [RP2.1] Panel, BioFire Diagnostics) or using an HadV-specific PCR on whole blood (Adenovirus ELITe MGB Kit, ELITech Group SpA) based on specific clinician request, which usually focuses on patients requiring critical care or in immunocompromised patients under the care of haematology/oncology teams. Testing on blood and eye samples was done using Adenovirus ELITe MGB® Kit, a clinically validated real-time PCR assay.

### Definitions of clinically significant hepatitis

AS-Hep-UA is defined as someone <16 years of age presenting no earlier than 1st October 2021 with an acute hepatitis and ALT or AST >500 IU/L, which cannot be accounted for by other causes (7). We additionally identified cases of AHUA in our dataset using a more relaxed definition, intending to capture cases that would not meet criteria for AS-Hep-UA. We defined AHUA as patients assigned diagnostic codes from the International Classification of Diseases 10th Revision consistent with hepatitis of an uncertain cause (ICD10) K759, K752, K720, K716, B178, B179, and B199 (**Table 2**) or patients with ALT>2x ULN. We also analysed data for presentations of diagnosed acute or chronic viral hepatitis A-E virus infection (B159, B162, B169, B180, B182, B171, B182, B172) as a baseline control, and to ensure these cases were excluded from the AHUA category.

**Table 2:** Diagnostic codes representing Acute Hepatitis of Unknown Aetiology (AHUA) and confirmed viral hepatitis A-E infection assigned to patients presenting as an emergency to Oxford University Hospitals NHS Foundation Trust (UK) between 2016 and 2022.

| ICD10 code | Condition(s) coded | Younger children<br>(18 months-6<br>years)<br>n=8 | Older children (7-15 years)<br>n=11 | Adults<br>(>15 years)<br>n=1,512 |
| --- | --- | --- | --- | --- |
| **Codes representing acute hepatitis of unknown aetiology (AHUA; n=407)** |  |  |  |  |
| K716 | Toxic liver disease with hepatitis, not elsewhere classified | <5 | <5 | 14 |
| K720 | Acute and subacute hepatic failure | <5 | <5 | 182 |
| K752 | Nonspecific reactive hepatitis | <5 | <5 | <5 |
| K759 | Inflammatory liver disease, unspecified | <5 | <5 | 97 |
| B178 | Other specified acute viral hepatitis | <5 | <5 | 5 |
| B179 | Acute viral hepatitis, unspecified | <5 | <5 | 71 |
| B199 | Unspecified viral hepatitis without hepatic coma | <5 | <5 | 23 |
| **Codes representing viral hepatitis A-E (n=1175)** |  |  |  |  |
| B159 | Hepatitis A | <5 | <5 | 32 |
| B162, B169, B180, B182 | Acute or chronic viral hepatitis B ± delta virus | <5 | <5 | 225 |
| B171, B182 | Acute or chronic viral hepatitis C | <5 | <5 | 860 |
| B172 | Acute viral hepatitis E | <5 | <5 | 26 |

### Data sharing

Anonymised LFT data were shared with the Summary Analysis of Laboratory Tests (‘SALT’) project (12), co-ordinated by the UKHSA as part of the UK-wide public health response, to contribute to a national picture of changes in the incidence of deranged liver function (for epidemiology and ongoing surveillance). The complete datasets analysed during the current study are not publicly available as they contain personal data but are available from the Infections in Oxfordshire Research Database (https://oxfordbrc.nihr.ac.uk/research-themes-overview/antimicrobial-resistance-and-modernising-microbiology/infections-in-oxfordshire-research-database-iord/), subject to an application and research proposal meeting the ethical and governance requirements of the Database. For further details on how to apply for access to the data and for a research proposal template please.

### Data analysis and statistical testing

Each presentation episode was considered independently; thus individuals may have featured more than once across the study duration. We used the first set of blood tests taken on presentation for analysis. An infecting pathogen was reported if at least one microbiology test was positive. Data were analysed using *R* v4.1.2 and visualised using *ggplot* v3.4.0. The code used for all analyses is hosted on GitHub (https://github.com/cednotsed/iORD_hepatitis.git). We tested for the presence of a non-monotonic trend using the non-parametric WAVK test (13), using its implementation in the *R* package *funtimes* (14). Fisher’s exact tests and Mann-Whitney U tests were performed using the *fisher.test* and *wilcox.test* functions in *R*. Odds ratios (OR) were calculated using conditional maximum likelihood estimation as part of the *fisher.test* function.

## RESULTS

### No evidence for an increase in hospital presentations or elevated liver enzymes during AS-Hep-UA outbreak

Data on 903,433 adults and children 18 months or older presenting acutely to OUH between 1^st^ March 2016 and 31^st^ December 2022 were analysed, representing 441,780 males and 461,632 females (and 21 individuals for whom sex was not recorded). The median age at presentation was 44 years (IQR 22-69 years), with 7.9%, 9.5%, and 82.6% classified as younger children, older children, and adults, respectively (**Table 1**). A median of 11,023 patients presented to the hospital per month, with a marked decline in the number of presentations in April 2020 coinciding with the implementation of SARS-CoV-2 (COVID-19) pandemic lockdown measures in the UK introduced on 26th March 2020 (**Supplementary Figure 1a**).

During the AS-Hep-UA outbreak, 1^st^ October 2021 – 31^st^ August 2022, minimal changes in the number of acute presentations per month was observed across any of the three age groups (all WAVK tests p>0.05; **Supplementary Figure 1a**). There was an overall increasing trend in the number of ALT tests requested for acutely presenting patients over time since March 2016 regardless of sex (*WAVK statistic* = 13.436, *p* < 0.0001; **Supplementary Figure 1b**). However, there was also an increasing trend in the number of ALT observations compared to WBC observations, which indicates increased ‘liver function scrutiny’ over time (**Supplementary Figure 1c**; *WAVK statistic* = 91.127, *p* < 0.0001).

Across the study duration, 59% of patient episodes had recorded blood tests. Among these, 90% had an ALT test and 1.2% an AST test. The median and IQR of ALT levels did not change significantly over time for any age group (**Figure 1a**; WAVK tests *p>*0.05), with no observable peak during the AS-Hep-UA outbreak. Similarly, the proportion of individuals with mild, moderate or severe derangement of ALT levels (1-2x, 2-5x and >5x ULN, respectively) remained relatively constant over time (**Figure 1b**; WAVK tests p>0.05). Therefore, there was no temporal association between elevated liver enzymes in patients presenting acutely to the hospital and the period of the AS-Hep-UA outbreak.

**Figure 1.**
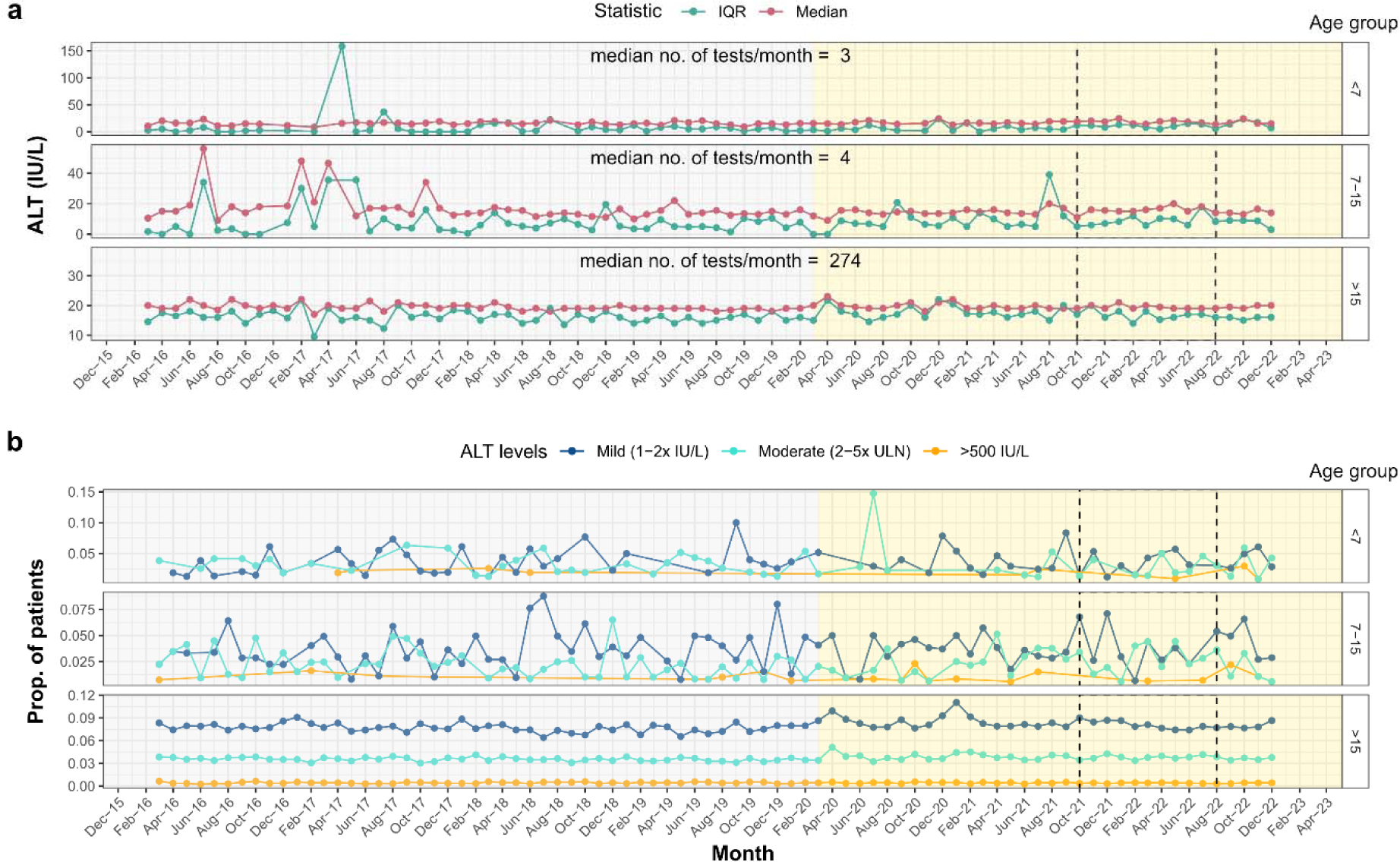
Temporal trends in ALT levels in patients presenting acutely to OUH. (a) median and IQR of ALT levels in patients presenting to OUH and (b) proportion of patients with mild, moderate and severe derangement in ALT levels aggregated at two month intervals. Relevant epochs are highlighted in grey (pre-COVID-19-pandemic), yellow (COVID-19 pandemic), and with dashed lines (start of AS-Hep-UA outbreak to end of first quarter of 2022).

### Increased incidence of AHUA in adults coinciding with AS-Hep-UA outbreak

We further investigated temporal trends based on ICD10 codes. Across the study duration, 1582 diagnostic codes representing AHUA or viral hepatitis A-E were assigned to 1531 distinct patient episodes (**Table 2**), of which 98% were adults (eight younger children, 11 older children, 1512 adults). The number of acute hepatitis diagnoses classified as AHUA or viral hepatitis A-E per month remained relatively constant from March 2016 to August 2018, with a peak between August 2018 to August 2019 followed by a decline from ∼23 diagnoses/month to ∼15-17 diagnoses/month from August 2019 to October 2021 (**Figure 2a**). Coinciding with the time interval of the AS-Hep-UA outbreak, an increasing trend in the 12-month moving average was observed between October 2021 and June 2022. This increasing trendline coinciding with the outbreak was seen primarily in ICD10 codes representing AHUA (**Table 2**), with the 12-month moving average of diagnoses per month doubling from five to ten between October 2021 to December 2022 (**Figure 2b**). The proportion of patients diagnosed with AHUA was higher during the AS-Hep-UA outbreak than outside this period (Fisher’s exact test p<0.0001; OR 3.01, 95% CI:2.20-4.12). Overall, these observations suggest an increased incidence of AHUA amongst adults during the AS-Hep-UA outbreak. We could not determine if this was the case for children, since only 2% of relevant diagnostic codes were assigned to children.

**Figure 2.**
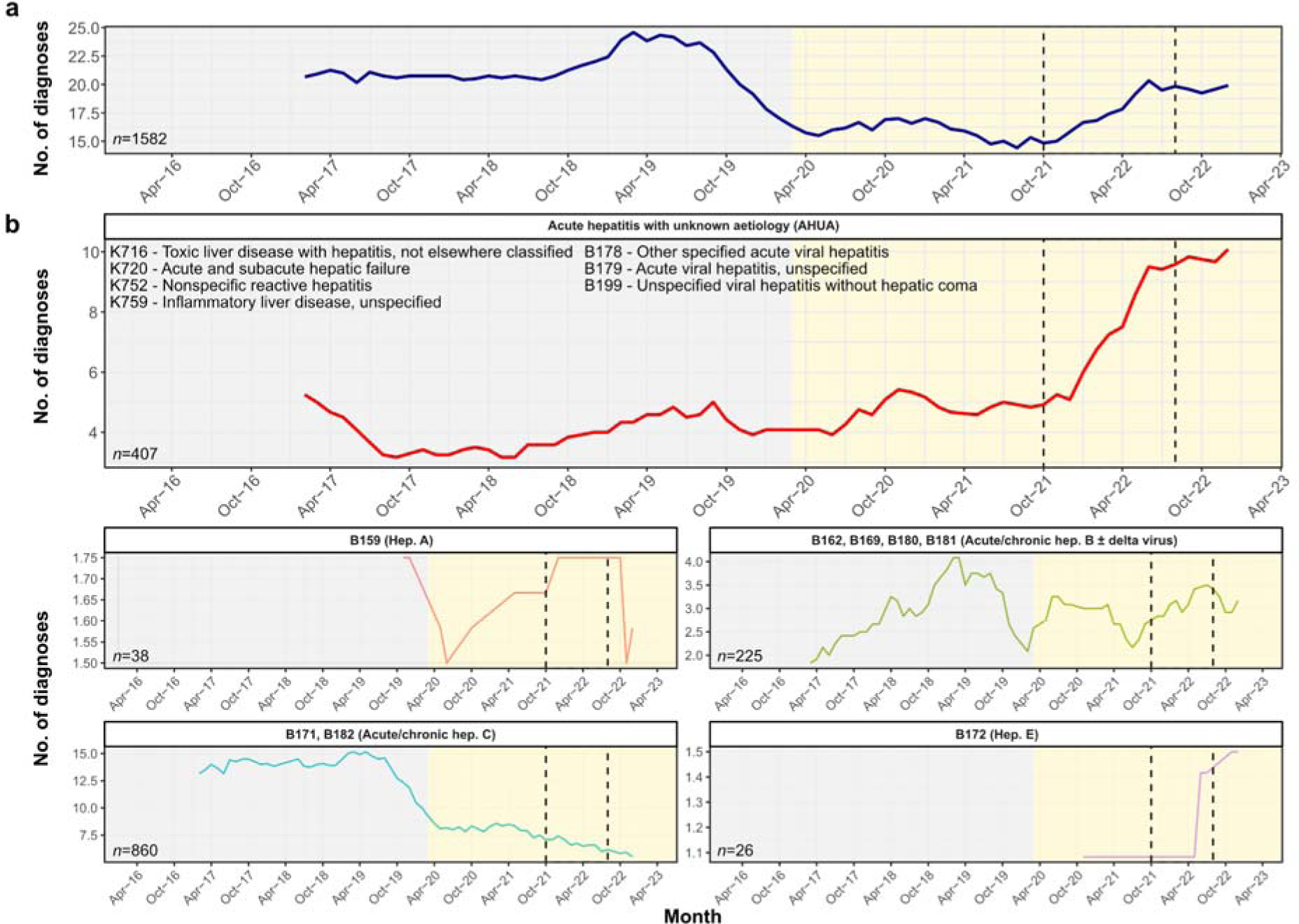
Temporal trends of acute hepatitis with unknown aetiology (AHUA) based on clinical coding at Oxford University Hospitals from 2016 to 2022. Twelve-month moving averages of (a) overall number of diagnoses (viral hepatitis A-E or AHUA) per month regardless of age group or sex, and (b), liver-related diagnoses with or without a specified causal agent. ICD10 codes and their described causal agents are annotated. B159, Hepatitis A virus infection without hepatic coma; B169, Acute hepatitis B virus infection without delta-agent and without hepatic coma; B171, Acute hepatitis C virus infection; B172, Acute hepatitis E virus infection; B181, chronic viral hepatitis B virus infection without delta agent; B182, chronic hepatitis C virus infection. Relevant epochs are highlighted in grey (pre-COVID-19-pandemic), yellow (COVID-19 pandemic), and with dashed lines (start of AS-Hep-UA outbreak to end of first quarter of 2022).

Compared to patients diagnosed with viral hepatitis A-E, those with AHUA had significantly higher ICU admission rates (both Fisher’s exact test p<0.0001; OR 5.01 within AS-Hep-UA outbreak and 3.90 outside outbreak) and longer hospitalisation periods (Mann-Whitney U tests p<0.05), both within and outside the AS-Hep-UA epoch. The mortality rate for the AHUA group was significantly higher than those diagnosed with viral hepatitis A-E outside of the AS-Hep-UA epoch (Fisher’s exact test p<0.0001; OR 19.8, 95% CI: 4.29-185), but not within the AS-Hep-UA epoch (p>0.05; OR=1.16, 95% CI: 0.0825-16.3). Additionally, compared to patients without AHUA, patients with AHUA also had significantly higher ICU admission (both Fisher’s exact tests p<0.0001; OR 41.7 within and 23.7 outside) and mortality rates (Fisher’s exact tests p=0.035 within and p<0.0001 outside; OR 6.99 within and 11.3 outside), and longer hospitalisation periods (both Mann-Whitney U tests p<0.0001; **Table 3**).

**Table 3:** **Outcomes of presentation to hospital among individuals presenting as an emergency to Oxford University Hospitals NHS Foundation Trust (UK) between 2016 and 2022**

| **Population** | **Hospital admission rate (%)** | **ICU admission rate (%)** | **Duration of hospital admission in hours (median, IQR)** | **Mortality rate (%)** |
| --- | --- | --- | --- | --- |
| All patients (n=903,433) | 30.9 | 1.3 | 51 (21-146) | 0.34 |
| HAdV tested (n=3707) | 78.5 | 18.2 | 101 (45-221) | 0.59 |
| HAdV untested (n=899,726) | 30.7 | 1.2 | 50 (21-145) | 0.34 |
| HAdV tested and positive (n=179) | 60.9 | 11.7 | 65 (31-139) | 0 |
| HAdV tested and negative (n=3528) | 79.4 | 18.5 | 102 (46-224) | 0.62 |
| Patients with diagnostic codes (n=1531) | 100 | 12.3 | 93 (35-234) | 1.1 |
| Patients with only diagnostic codes indicative of AHUA (n=391) | 100 | 26.1 | 146 (57-333) | 3.3 |
| Patients with diagnostic codes indicating viral hepatitis A-E (n=1140) | 100 | 7.5 | 79 (29-199) | 0.35 |
| Patients without AHUA (903,042) | 30.9 | 1.3 | 50 (21-145) | 0.34 |

### Increased incidence of *HAdV* infections during AS-Hep-UA outbreak but not associated with deranged liver enzymes or poorer patient outcomes

Across the study duration, we retrieved 3707 distinct patient records that included microbiology tests for HAdV infection, of which 179 were positive (4.8%). The positivity rate was highest in younger children, among whom 124/781 (15.9%) of HAdV tests were positive, compared to older children (9/440, 2.0% positive) and adults (46/2486, 1.9% positive), in keeping with the known epidemiology of HAdV infection (15–17). None of the HAdV-infected patients were given ICD10 codes indicative of AHUA across the study duration. A minority (16/179; 9%) of HadV positive results were derived from eye swabs, which is unlikely to have influenced any overall trends.

There was an increase in the number of HAdV-tests undertaken between April 2021 and April 2022 (**Figure 3a**), and a significantly higher number of HAdV tests performed relative to all microbiology tests performed during the AS-Hep-UA epoch (Fisher’s exact test p<0.0001; OR 1.27, 95% CI:1.17-1.37). These findings indicate increased clinician scrutiny for HAdV coinciding with the AS-Hep-UA outbreak. The proportion of HAdV-positive tests during the AS-Hep-UA epoch was significantly higher than outside of the AS-Hep-UA epoch at 60/839 (7.2%) vs 119/2868 (4.1%) respectively; (Fisher’s exact test *p*<0.001; OR 1.78, 95% CI:1.27-2.47). Additionally, there was an increase in the incidence and proportion of HAdV-positive tests in younger children during the AS-Hep-UA outbreak relative to the period preceding the outbreak (**Figure 3b**). However, there were also multiple peaks in the proportion of HAdV-positive tests across the entire study duration (**Figure 3b**), indicating previous periods of high HAdV-positivity before the AS-Hep-UA epoch. These results indicate that despite an increased incidence of HAdV infections during the AS-Hep-UA outbreak, this was not an unusual aspect of the local HAdV epidemiology, and could have been partly accounted for by increased clinician scrutiny for HAdV in the same period.

**Figure 3.**
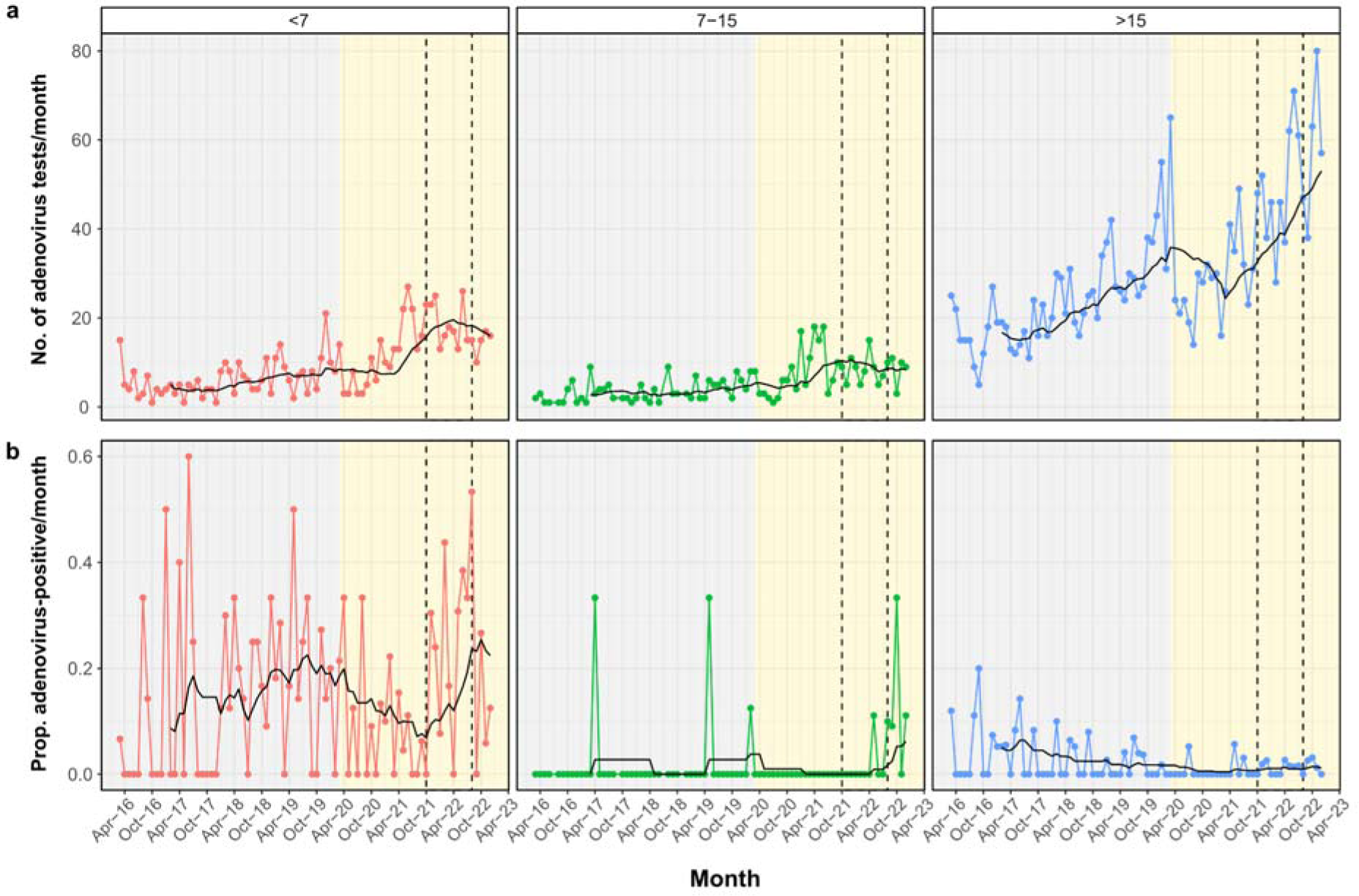
Temporal trends of HAdV-related microbiological tests requested at OUH from 2016 to 2022. (a) Number of HAdV tests requested and (b) the proportion of HadV-positive tests per month. Relevant epochs are highlighted in grey (pre-COVID-19-pandemic), yellow (COVID-19 pandemic), and with dashed lines (start of AS-Hep-UA outbreak to end of first quarter of 2022). Red, green, blue and black lines show the (a) number of HAdV tests or (b) proportion of HAdV-positive tests per month for younger children (<7 years), older children (7-15 years), adults (>15), and 12-month simple moving average, respectively.

The proportion of patients with mild, moderate or severe derangement of ALT, AST, bilirubin, GGT, CRP or WBC did not differ significantly between those testing positive vs. negative for HAdV (Fisher’s exact test p>0.05; **Supplementary Figure 2**). The proportion of patients with low albumin was significantly smaller for those testing HAdV-positive vs. negative (Fisher’s exact test p=0.01; **Supplementary Figure 2**). However, a significantly higher proportion of HAdV-positive patients had mild derangement of ALP compared to those with no HAdV diagnosis (Fisher’s exact test p=0.004; **Supplementary Figure 2**). A similar association with raised ALP was also present in confirmed rhinovirus/enterovirus infections (Fisher’s exact test p=0.004), indicating that mild derangement of ALP is not unique to HAdV infections (data not shown).

HAdV testing focuses primarily on a clinically vulnerable group, shown by higher rates of hospital admission, ICU admission and mortality among those receiving a HAdV test (irrespective of the test result) compared to the untested population (78.5% vs 30.7%, 18.2% vs 1.2%, and 0.59% vs 0.34%, respectively; **Table 3**). The HAdV-positive group fared somewhat better than those who tested negative, with lower hospital admission (both Fisher’s exact tests p<0.001) and significantly shorter hospital stays (both Mann-Whitney U tests p<0.05), whether within or outside the AS-Hep-UA epoch. Additionally, the HAdV-positive group had significantly lower ICU admission rates within the AS-Hep-UA epoch, but not outside the epoch (Fisher’s exact tests p=0.015 and p=0.35, respectively) (data not shown). No HAdV-positive patients died across the entire study duration. Characteristics of the population testing positive for HAdV are presented in **Table 4**.

**Table 4:** Characteristics of 179 patient episodes with HAdV-positive microbiology tests among children and adults presenting as an emergency to Oxford University Hospitals NHS Foundation Trust (UK) between 2016 and 2022.

| Characteristic |  | Younger children<br>(18 months-6<br>years) | Older children (7-15 years) | Adults<br>(>15 years) |
| --- | --- | --- | --- | --- |
| Total number tested for HAdV infection |  | 781 | 440 | 2486 |
| Total number positive for HAdV infection<br>(% of those tested) |  | 124/781 (15.9%) | 9/440 (2.0%) | 46/2486 (1.9%) |
| Epoch of<br>presentation<sup>a</sup> | Pre-COVID-19<br>(n=79 HAdV-positive) | 45/79 (57.0%) | 4/79 (5.1%) | 30/79 (38.0%) |
|  | COVID-19 pandemic<br>(n=100 HAdV-positive) | 79/100 (79.0%) | 5/100 (5.0%) | 16/100 (16.0%) |
|  | AS-Hep-UA (n=60<br>HAdV-positive) | 51/60 (85.0%) | 2/60 (3.3%) | 7/60 (11.7%) |
| Levels of<br>derangement for<br>ALT | Not tested | 66 | 3 | 10 |
|  | Normal | 53 | 5 | 25 |
|  | Mild<br>(Up to 2x ULN) | <5 | 0 | 7 |
|  | Moderate<br>(2-5x ULN) | <5 | <5 | <5 |
|  | Severe<br>(>5x ULN) | 0 | 0 | <5 |
| Inflammatory<br>markers (median,<br>IQR) | CRP (mg/L) | 47.2 (10.6-73.3) | 24.0 (11.8-88.6) | 81.9 (23.3-143.7) |
|  | White cells (x10<sup>9</sup>/L) | 11.6 (8.53-17.7) | 9.1 (6.60-9.61) | 10.4 (6.73-14.6) |
| Admission rate |  | 74/124 (59.7%) | 5/9 (55.6%) | 30/46 (65.2%) |
| ICU admission rate |  | 9/124 (7.3%) | 0/9 (0%) | 12/46 (26.1%) |
| **Duration of hospital admission in hours (median, IQR)** |  | 56 (26-95) | 118 (33-309) | 125 (49-450) |
| **Death rate during admission** |  | 0/124 (0%) | 0/9 (0%) | 0/46 (0%) |
| **Vital signs at presentation (median, IQR)** | Heart rate | 134 (112-145) | 111 (109-123) | 92 (84-106) |
|  | Diastolic blood pressure | 61 (54.0-71.5) | 81.5 (74.0, 86.3) | 73.0 (61.3-82.0) |
|  | Systolic blood pressure | 104.0 (94.5-110.0) | 109.0 (105.0-116.8) | 123.5 (113.5-135.8) |
|  | Tympanic temperature | 37.5 (36.7-38.1) | 37.3 (36.9-37.9) | 36.9 (36.4-37.6) |
|  | Oxygen saturation | 98.0 (96.0-99.3) | 98.0 (98.0-100.0) | 96.0 (95.0-98.3) |
|  | Respiratory rate | 28.0 (24.0-35.0) | 27.5 (22.5-30.5) | 19.0 (17.0-21.0) |

<sup>a</sup> AS-Hep-UA epoch is nested within the Covid-19 pandemic period. Epochs were considered as pre-COVID-19 (1st March 2016 - 10 March 2020), COVID-19 pandemic period (11th March 2020 - 31st December 2022), and the AS-Hep-UA outbreak (1st Oct 2021 - 31 Aug 2022).

## DISCUSSION

Anonymised clinical data from EHRs offers access to large datasets, providing power in numbers to determine overall trends reflecting clinical epidemiology and its influence on morbidity, mortality and health service workload. Following an outbreak of AS-Hep-UA in children during 2022, we reviewed anonymised EHRs to determine whether we could identify evidence of the outbreak in the population of Oxfordshire, UK, presenting acutely to hospital, reviewing incidence of elevated liver enzymes, changes in presentations based on clinical coding and/or an increase in incidence of HAdV infection. We identified an increased incidence of episodes coded as AHUA in adults and an increased incidence of HAdV infections in younger children coinciding with the AS-Hep-UA outbreak. While the latter may be partially accounted for by increased clinician scrutiny during this period, the pattern was not observed to the same level in older children, and not at all in adults despite similar increases in clinical scrutiny for all age groups.

Although we have captured data for a large population over a period of almost seven years, this remains a small data set within which to identify rare events, and our region was not known to be directly affected by the AS-Hep-UA outbreak. There was no evidence for increased incidence of abnormal liver enzymes in children and adults, nor associations between HAdV infections and elevated liver transaminases. Some of the AHUA reported in the intensive care population may be related to liver ischaemia from diverse causes associated with critical illness, rather than liver-specific pathology.

Among patients presenting acutely for hospital-based care, HAdV testing is biased towards a vulnerable group. Even in this high-risk population, those testing positive for HAdV had lower admission rates than those testing negative, reinforcing the view that this virus is generally benign and self-limiting, with a low risk of serious complications. The lack of associations between HAdV infections and deranged liver enzymes is concordant with the fact that HAdV infections typically lead to mild respiratory or gastrointestinal disease, and that hepatitis is an unusual complication (17). Co-infections with HAdV and other viruses such as respiratory syncytial virus (RSV) have been linked to poorer outcomes (17), but the small number of HAdV infections identified in this study precluded robust analysis of mixed infections.

Monitoring of EHRs may be an effective and low-cost surveillance tool that allows identification of trends that could be of concern - e.g. deranged laboratory parameters and/or changes in recorded diagnoses based on microbiology tests or coding. Such strategies could potentially be developed to provide an ‘early warning’ system to allow clinical and public health authorities to review data in real time, cross-compare between regions, pick up possible outbreaks, and implement enhanced surveillance and public-health messaging if necessary. However, as we have demonstrated, there is a need to consider which populations and patient groups are represented, and to account for changes in the behaviour of healthcare practitioners over time, which can account for increased testing (and thus dilute the proportion of those testing positive for a rare event), or alternatively enhance detection of minor abnormalities which are not clinically relevant.

There are various caveats to our approach for detection of AHUA. If there were milder cases of disease in the population during or preceding the documented AS-Hep-UA outbreak, these may not have presented to hospital at all. Even among those presenting to healthcare, many patients did not have liver enzymes measured. Thus, the IORD dataset provides only a limited view of the whole population, which is not true community surveillance. Hospital admission data are also biased by repeated representation of the same individuals, and over-represent populations who preferentially present to emergency care rather than accessing primary care. Data for individuals presenting to hospital through different routes (for example, a transfer between hospitals or a primary care referral that bypassed the emergency department) will not have been captured. Clinical datasets are always subject to missingness, which may not be random. Detection of relevant pathogens also depends on the sample type collected, as relevant viruses may be variably identified from blood, respiratory samples, urine, stool or tissue. Our analyses also relies on consistent clinical coding of patient episodes, which has been shown to be subject to some inaccuracy and/or variability (18).

We now know that the severe clinical outcomes during the AS-Hep-UA outbreak in children was likely driven by AAV2 as the leading aetiological agent. However, AAV2 requires co-infection with a ‘helper’ virus (including, but not limited to, HAdV) to replicate, and this appears to be a requirement for the development of liver pathology (3–5). Retrospective analysis of EHRs cannot be used to investigate the epidemiology of AAV infection, as these viruses are not part of clinical diagnostic testing pathways and have only been identified as an agent of AS-Hep-UA through retrospective metagenomic sequencing (3–5). Additionally, HAdV infection is typically screened only in individuals presenting with specific pathology, or in those at high risk for complications of infection due to immunocompromise or other underlying health conditions. The specific subtype of HAdV implicated in the outbreak, 41F, is not routinely discriminated from other types by clinical testing. Other possible ‘helper viruses’ include human herpesviruses (5), many of which are ubiquitous in the population and characterised by long-term carriage and latency, making it difficult to distinguish between clinically relevant episodes and subclinical reactivation. Thus routinely-collected clinical datasets do not include screening for all relevant pathogens, and detection of implicated pathogens can be difficult to interpret due to the detection of commensal or bystander organisms. Overall, wider adoption of metagenomics-based diagnostics has the potential to further enhance the utility of EHRs to investigate future outbreaks but interpretation is complex (4).

Longer-term collation of data from multiple regions would offer a more powerful approach that could be extended to other diseases. However, surveillance through EHRs requires the establishment of suitable and systematic data-processing infrastructure and governance frameworks, in addition to investment of personnel and resources, if it is to become a real-world surveillance tool.

## Data Availability

The complete datasets analysed during the current study are not publicly available as they contain personal data but are available from the Infections in Oxfordshire Research Database (https://oxfordbrc.nihr.ac.uk/research-themes-overview/antimicrobial-resistance-and-modernising-microbiology/infections-in-oxfordshire-research-database-iord/), subject to an application and research proposal meeting the ethical and governance requirements of the Database. For further details on how to apply for access to the data and for a research proposal template please.

## SUPPLEMENTARY FIGURES

**Supplementary Figure 1.**
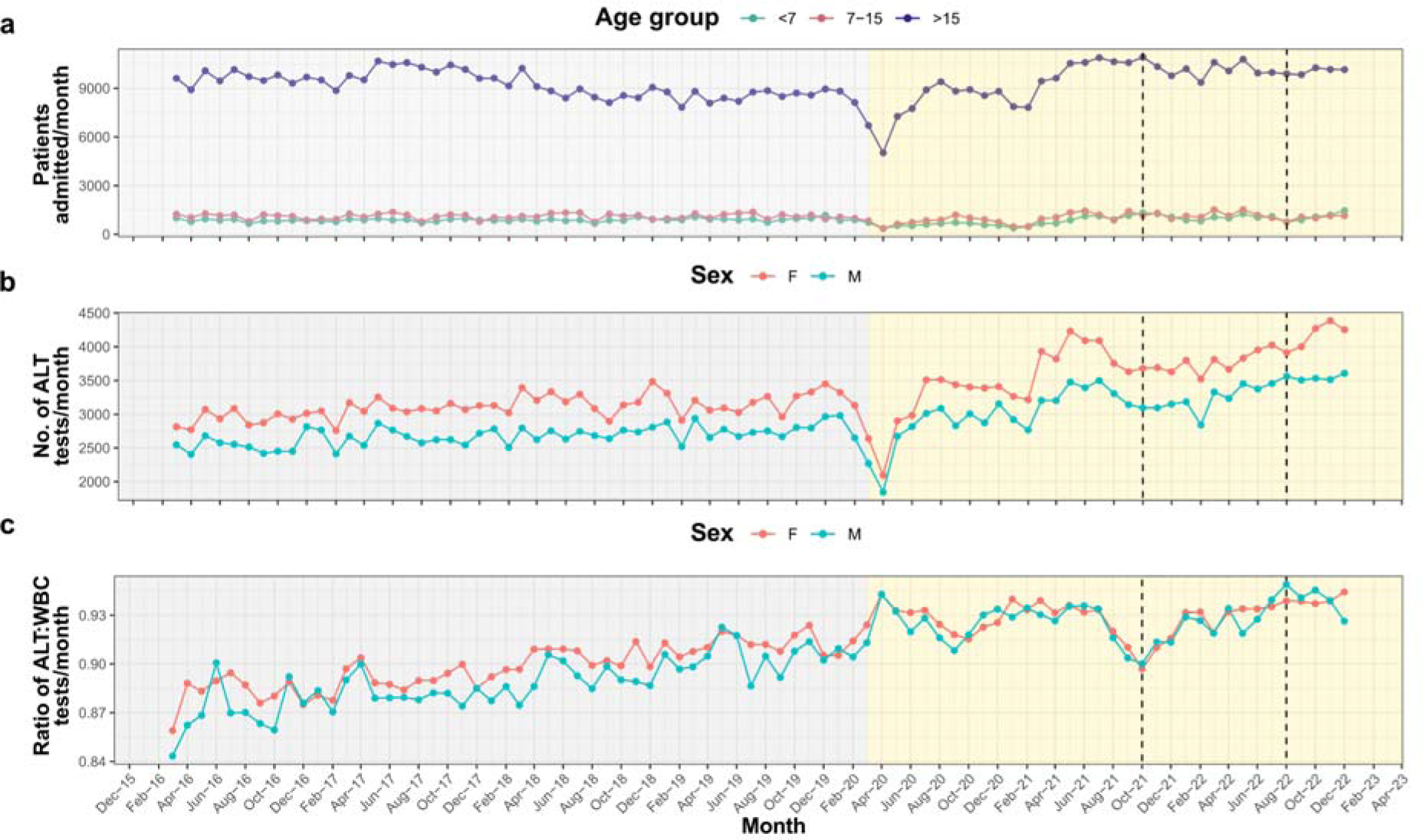
Temporal trends in presentations to hospital and laboratory requests per month over the time period of the study. (a) Number of patients presenting to hospital, (b) number of ALT tests requested, and (c) ratio of ALT to WBC tests requested. Relevant epochs are highlighted in grey (pre-COVID-19-pandemic), yellow (COVID-19 pandemic), and with dashed lines (start and end of AS-Hep-UA outbreak to end of first quarter of 2022).

**Supplementary Figure 2.**
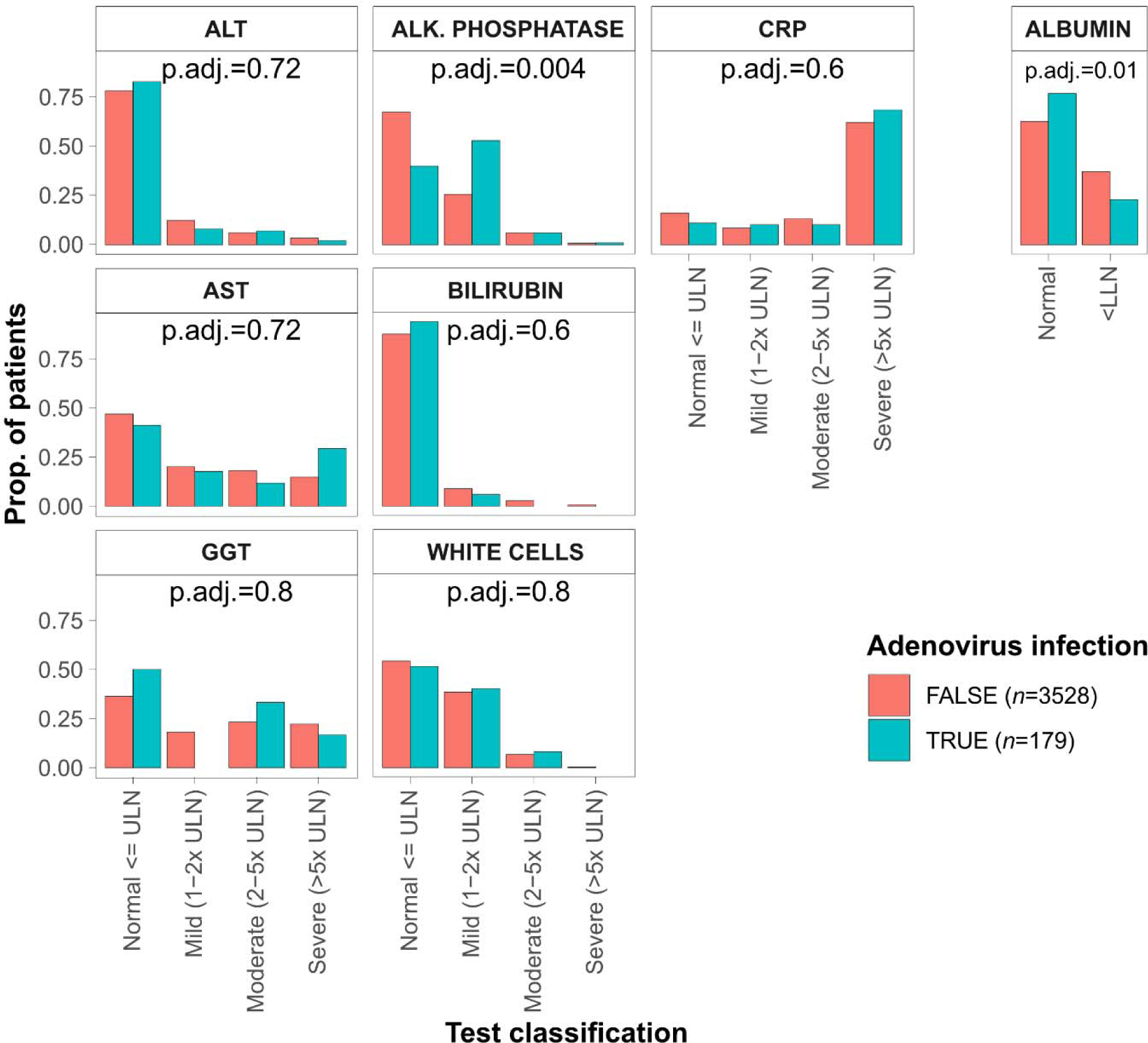
Associations between HAdV infection and blood biomarkers. Barchart showing the proportion of patients, stratified by HAdV infection, with various levels of derangement of blood biomarkers. Albumin levels were considered deranged if they were less than the lower limit of normal (32g/L). For each blood biomarker, Fisher’s exact test was used to determine if the proportion of patients falling into each derangement category differed significantly between patients with HAdV infections or otherwise. Benjamini-Hochberg procedure was used to correct for multiple testing and adjusted p-values, where available, were annotated.

